# SARS-CoV-2 Screening Among Symptom-Free Healthcare Workers

**DOI:** 10.1101/2020.07.31.20166066

**Authors:** Ryan T. Demmer, Angela K. Ulrich, Talia D. Wiggen, Ali Strickland, Brianna M. Naumchik, Shalini Kulasingam, Steven D. Stovitz, Clarisse Marotz, Pedro Belda-Ferre, Greg Humphrey, Peter De Hoff, Louise Laurent, Susan Kline, Rob Knight

## Abstract

**Background:** Transmission of severe acute respiratory syndrome coronavirus 2 (SARS-CoV-2) is possible among symptom-free individuals and some patients are avoiding medically necessary healthcare visits for fear of becoming infected in the healthcare setting. Limited data are available on the point prevalence of SARS-CoV-2 infection in symptom-free U.S. healthcare workers (HCW).

**Methods:** A cross-sectional convenience sample of symptom-free HCWs from the metropolitan area surrounding Minneapolis and St. Paul, Minnesota was enrolled between April 20^th^ and June 24^th^, 2020. A participant self-collected nasopharyngeal swab (NPS) was obtained. SARS-CoV-2 infection was assessed via polymerase chain reaction. Participants were queried about their willingness to repeat a self-collection NPS for diagnostic purposes. We had >95% power to detect at least one positive test if the true underlying prevalence of SARS-CoV2 was ≥1%.

**Results:** Among n=489 participants 80% were female and mean age±SD was 41±11. Participants reported being physicians (14%), nurse practitioners (8%), physician’s assistants (4%), nurses (51%), medics (3%), or other which predominantly included laboratory technicians and administrative roles (22%). Exposure to a known/suspected COVID-19 case in the 14 days prior to enrollment was reported in 40% of participants. SARS-CoV-2 was not detected in any participant. Over 95% of participants reported a willingness to repeat a self-collected NP swab in the future.

**Conclusions:** The point prevalence of SARS-CoV-2 infection was likely <1% in a convenience sample of symptom-free Minnesota healthcare workers from April 20^th^ and June 24^th^, 2020. Self-collected NP swabs are well-tolerated and a viable alternative to provider-collected swabs to preserve PPE.

## INTRODUCTION

Current evidence suggests that nearly half of new severe acute respiratory syndrome coronavirus 2 (SARS-CoV-2) infections are due to transmission from asymptomatic or presymptomatic (i.e. symptom-free) individuals.^1-4^

Healthcare workers (HCW) have an increased risk of SARS-CoV-2 infection given their exposure to the virus while serving on the frontlines of the COVID-19 pandemic. HCW can inadvertently be a source of transmission since close contact with patients is often required for effective health care. However, it is also possible that the risk of infection among HCW might be similar to community risk as was recently seen in New York.^5^

To date there are limited data on the point prevalence – an absolute measure of infection – of infection among symptom-free HCW. These data can inform the potential role HCW play in transmission, guide testing recommendations and inform infection modeling studies. In addition, given the evidence that patients are avoiding medically necessary healthcare visits for fear of becoming infected, absolute measures of infection can inform the potential risk that a patient might encounter a symptom-free healthcare professional.^6^

To address this question, we screened symptom-free HCW for SARS-CoV-2. Additionally, to preserve personal protective equipment (PPE), we implemented a protocol for self-collection of nasopharyngeal swabs (NPS) and surveyed participants about their perceived quality of a self-collected vs. provider collected NPS.

## METHODS

### Participants

A convenience sample of individuals working in Minnesota healthcare facilities located in the Minneapolis/St. Paul metropolitan area was enrolled. Participants were identified via social media advertisements and enrolled from April 20^th^–June, 24^th^, 2020. Eligibility criteria were: i) employed or volunteering in a healthcare facility; ii) free of fever, chills, anosmia, pharyngitis, recently developed persistent cough, nasal congestion suspected to be unrelated to season allergies; iii) age 18-80 years; iv) not pregnant. A total of 489 participants provided self-collected NPS (Figure 1). The study was approved by the University of Minnesota Institutional Review Board. All participants provided informed consent.

**Figure 1.**
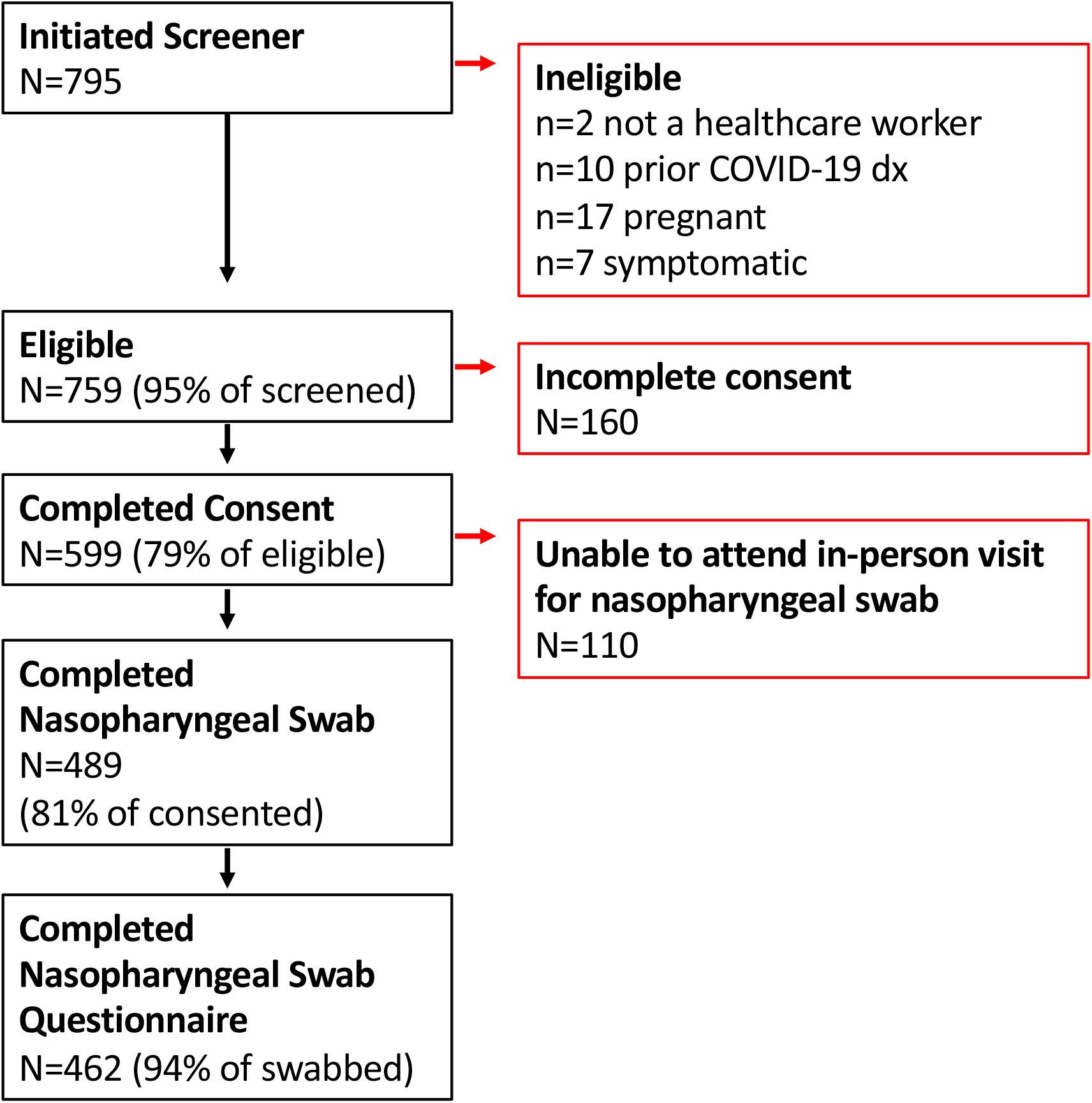
Participant Flow Diagram.

### Self-Collected Nasopharyngeal Swab (NPS)

Participants advanced a nylon flocked-tip NPS through the nasal passage (bilaterally) 3-4 inches into the nasopharynx and swirled the swab 360 degrees for 5 seconds. The swab tip was preserved in 3 mL of 95% ethanol, immediately placed on an ice bath, and transferred to a −80°C freezer. Samples were shipped overnight on dry ice to UC San Diego.

### Laboratory Assessment of SARS-CoV-2

Samples were processed within 48 hours of receipt at UC San Diego. Total nucleic acid was extracted from the swab heads using the MagMAX™ Microbiome Ultra Nucleic Acid Isolation Kit (A42357) and eluted in 100 µl nuclease-free H2O. SARS-CoV-2 Screening was performed using the one-step Applied Biosystems TaqPath COVID-19 Combo Kit (A47814) (https://assets.thermofisher.com/TFS-Assets/LSG/manuals/MAN0019181_TaqPath_COVID-19_IFU_EUA.pdf) following the manufacture’s protocol with the following exceptions: The reaction volume was scaled down to 3 µL with proportional reagent scaling and a replacement of ∼94% of the water with participant RNA. Additionally, the MS2 phage spike-in control was diluted 160-fold to improve sensitivity through reducing competition for reagent material within the multiplex RT-qPCR reaction. Samples were prepared in 384 well reaction plates using a mosquito® HV Robotic Liquid Handler (SPT Labtech) and a mosquito® X1 (HV) Robotic Liquid Handler (SPT Labtech). The RT-qPCR was analyzed in a QuantStudio5 qPCR instrument (ThermoFisher Scientific). Positive controls for each SARS-CoV-2 target amplified as expected, as well as all MS2 sample controls. None of the negative controls amplified.

### Questionnaires

Prior to the NPS, participants completed online surveys. After the NPS, participants were queried about their perception of the procedure relative to NPS they have performed on patients. They reported their level of discomfort with the self-swab on a scale of 1 (no discomfort) to 10 (the most discomfort they have ever experienced), their likelihood of repeating a self-collected NPS for clinical or research purposes.

### Statistical analysis

Analyses were preformed using SAS version 9.4. Descriptive characteristics are reported as mean±SD for continuous variables and %(n) for categorical variables. Bivariate analyses, t-tests and Chi-Square tests assessed statistical significance.

We had >95% power to detect at least one positive test if the true underlying prevalence of SARS-CoV2 was ≥1%.

## RESULTS

Among 489 participants enrolled, the mean age was 41±11 and 80% were female. All participants worked in facilities located in the seven-county Minneapolis and St. Paul metropolitan area. The average number of people living with participants was 2±1.4 and 12% reported living alone. The average number of children living with participants was 0.9±1.1 and 50% reported having at least one child at home.

The average time between NPS collection and laboratory testing was 36±18 (range=2-68) days. SARS-CoV-2 was not detected in any sample.

In the 14 days prior to enrollment, 40% of participants reported a known COVID-19 exposure. This proportion varied by venue (p<0.0001, Table 1) and role (p<0.01, Table 1). PPE use was high with only 1.4% of participants reporting no PPE use and this occurred among individuals without patient contact.

**Table 1.**
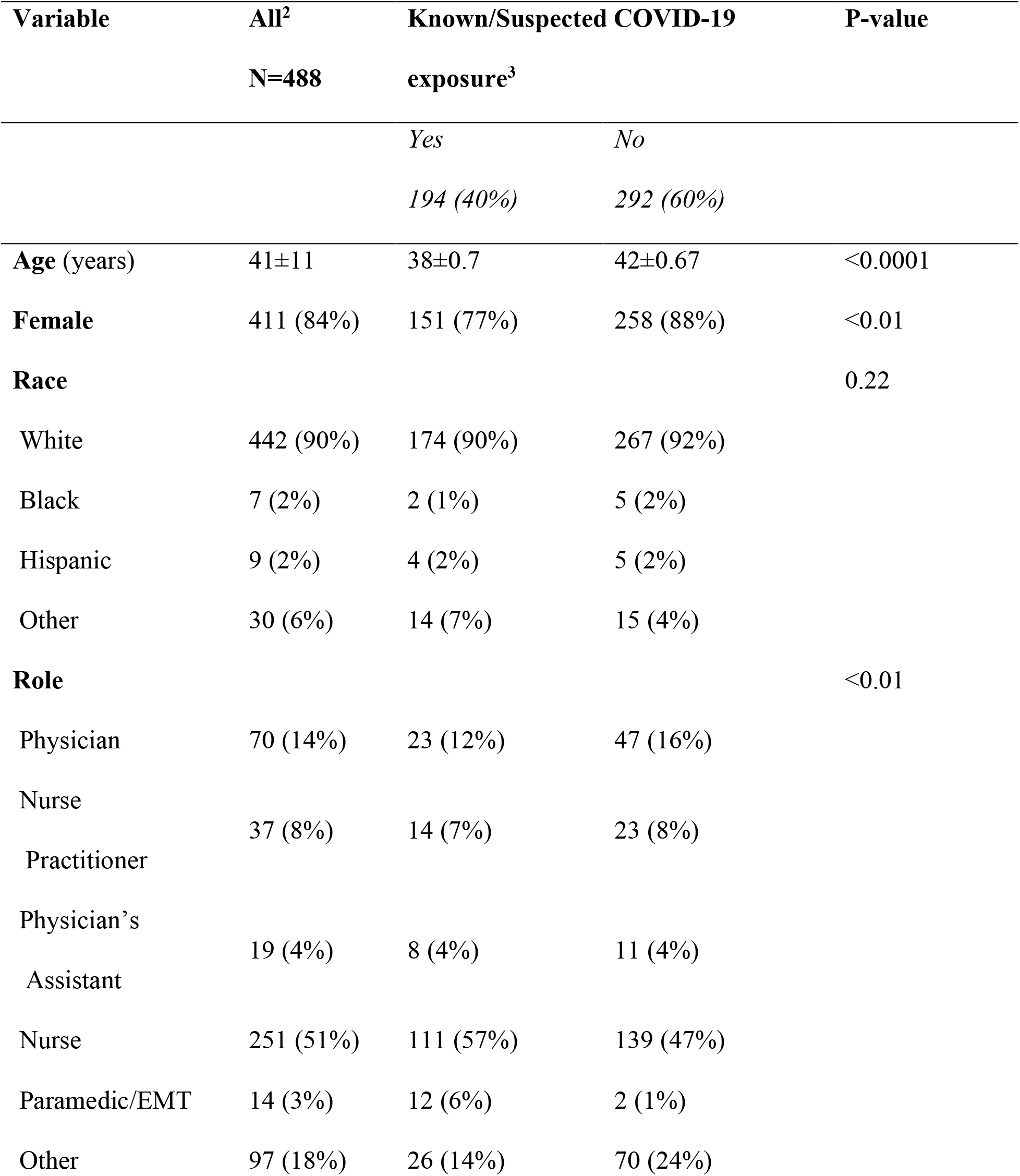
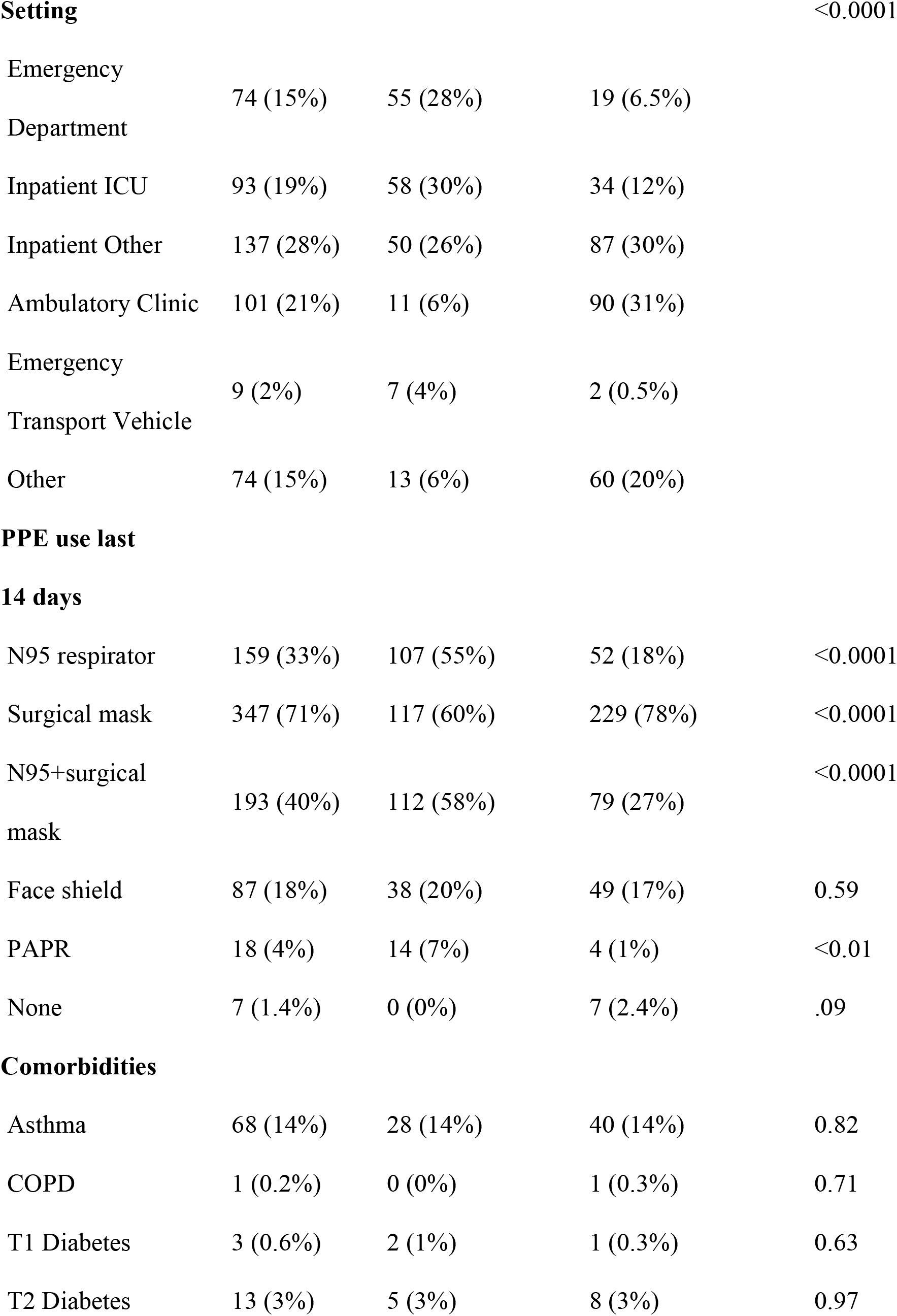
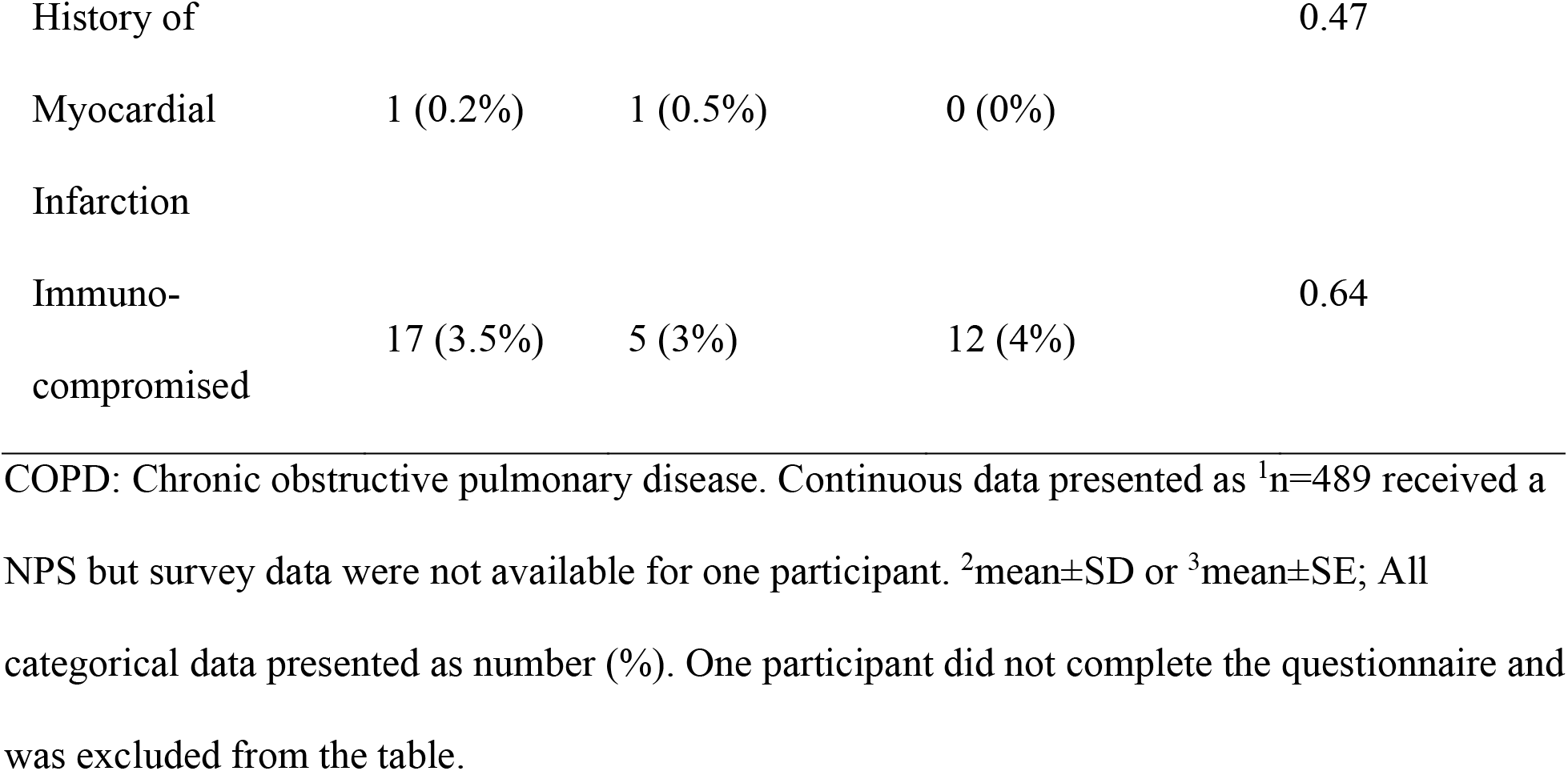
General characteristics of n=488^1^ Minnesota healthcare workers according to COVID-19 exposures within 14 days preceding enrollment. Enrollment occurred between April 20^th^, 2020 and June 24^th^, 2020.

The mean score for discomfort related to the self-collected NPS was 4.5±2.0 (range=1-10, Figure 2). Among the 62% (n=287) of participants who reported performing an NPS on a patient, 89% indicated that their self-swabbing depth was ≥ the depth of prior patient swabs, and 95% reported that their self-swab was ≥ the duration of previous patient swabs. Over 95% of participants reported a willingness to repeat a self-collected NPS in the future for either clinical or research purposes (Figure 2); 24% preferred a provider collected-swab, 57% preferred self-collection and 19% reported no preference.

**Figure 2.**
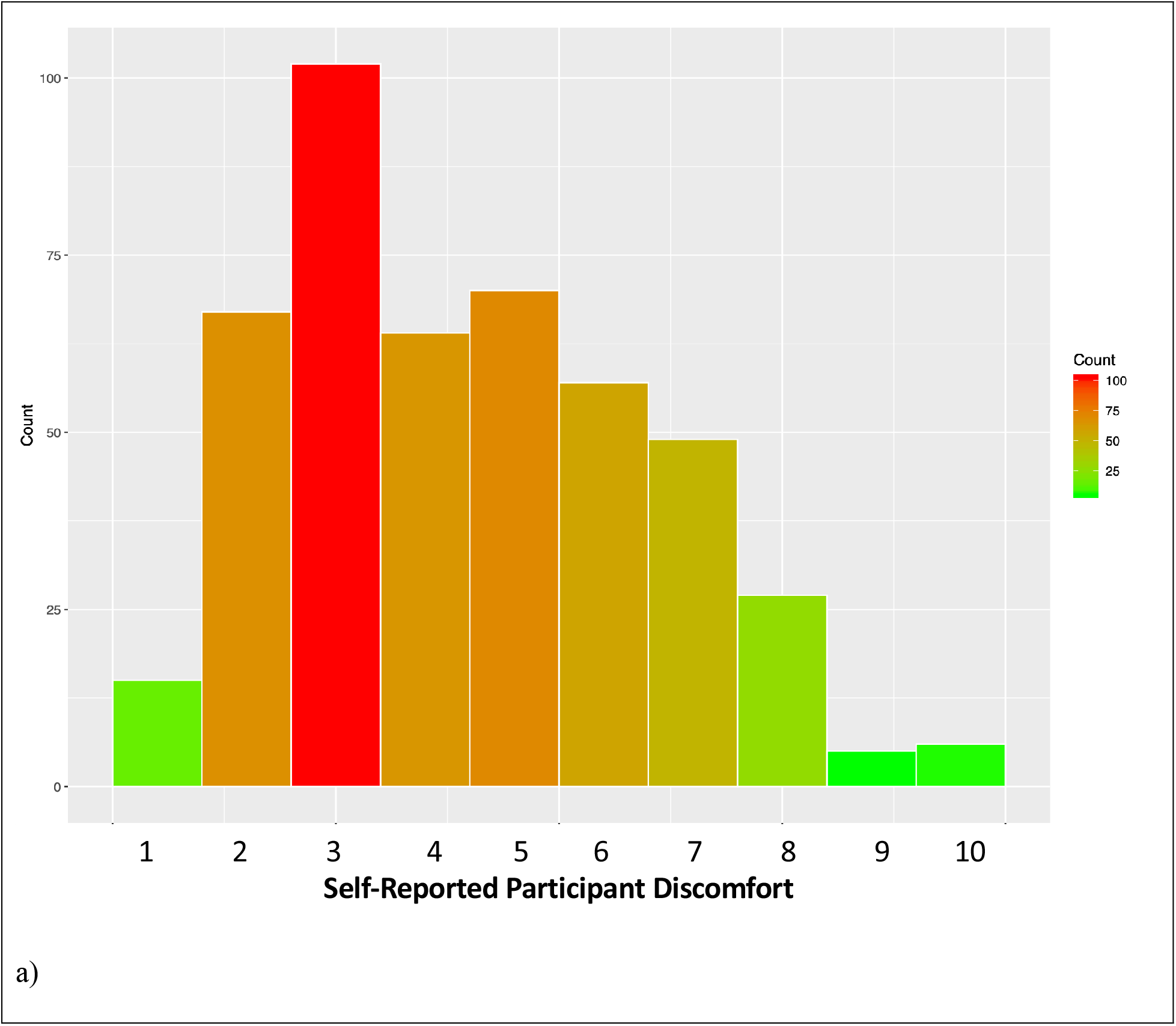
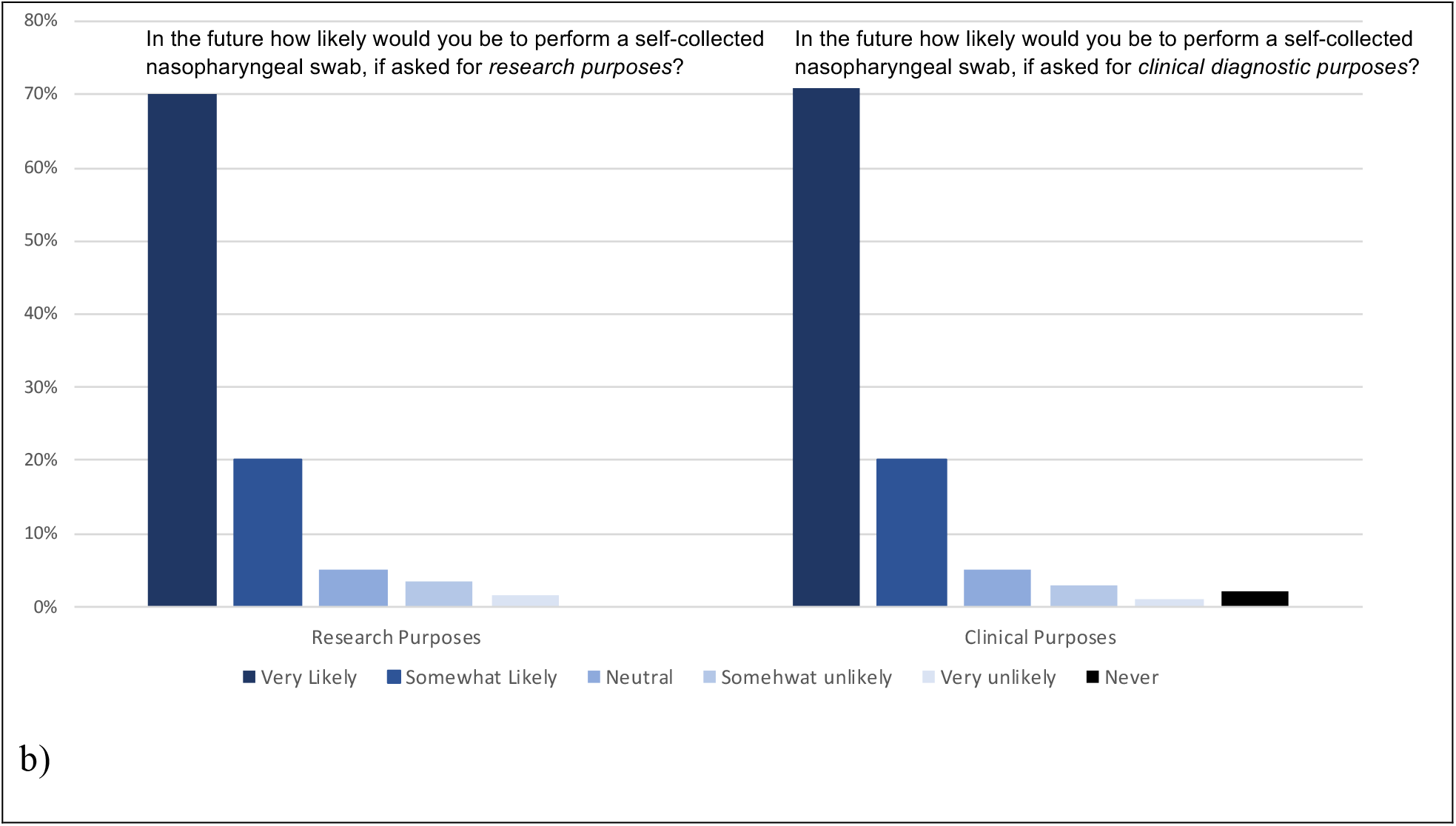
Participant perceptions of a self-collected nasopharyngeal swab. a) Histogram showing the distribution of responses to the following question: ‘On a scale of 1 to 10 how much discomfort did you experience during your self-swab (1 = no discomfort and 10 = the most discomfort you have ever experienced)?’; b) response patterns to questions about the future likelihood of performing a self-collected nasopharyngeal swab for research vs. clinical diagnostic purposes.

## DISCUSSION

We did not detect any SARS-CoV-2 positive individuals among a convenience sample of symptom-free HCW. Based on our power calculations, this strongly suggests that the point prevalence of SARS-CoV-2 in our study sample of symptom-free HCW was <1%. This is consistent with results in the U.S. population and in MN specifically during the period these samples were collected. National seroprevalence estimates reported by the Centers for Disease Control, including Minnesota, ranged from 1%-7%, and the estimate from Minnesota during the period from April 20th – May 12th, 2020, was 2.2%^7^. Seroprevalence estimates reflect an approximation of SARS-CoV-2 cumulative incidence since the beginning of the pandemic and are expected to be higher than the point prevalence estimates as the pandemic progresses. Low SARS-CoV-2 point prevalence in HCWs, despite increased relative risk for infection compared to the general population^8^, is plausible since HCW are prioritized to receive PPE and are trained in infection control which likely translates into reduced infection risk.

Our findings from the self-collected NPS survey suggest that the self-collection of NPS was acceptable to HCW, and that the depth and duration of swabbing in the nasopharynx is consistent with provider performed swabbing. Broader implementation of self-collection protocols could preserve PPE and enhance testing capacity as the pandemic progresses.

The sensitivity of our screening tests might have been low due to the use self-collected NPS, although recent studies report self-collection protocols to have acceptable sensitivity^9,10^. Tests among symptom-free individuals could also have reduced sensitivity, however, prior studies in asymptomatic pregnant women^11,12^ and residents of long-term care facilities^13^ have detected high SARS-CoV-2 prevalence. As this was a convenience sample, it is not representative of all HCWs in Minnesota, nor is it representative of what future SARS-CoV-2 prevalence estimates might be among symptom-free HCW in settings with high community prevalence. Importantly, there was no systematic inclusion of participants from long-term care facilities where infection risk has been notably higher in Minnesota.

Our results suggest that while the healthcare worker force is very likely to be at increased relative risk for infection compared to the general population^8^, the point prevalence of SARS-CoV-2 infection was low in symptom-free Minnesota healthcare workers. If true, the probability of encountering an infected symptom-free healthcare professional during a medically necessary healthcare visit is likely low when community point prevalence is low. Self-collected NP swabs are acceptable to participants and might be a future alternative to provider-collected swabs to preserve PPE. Ongoing monitoring of infection in healthcare workers will be important as the pandemic progresses and community transmission rises across the country.

## Data Availability

De-identified data can be made available upon request to Dr. Demmer.

## Thank you

We are also profoundly grateful for the study participants who have donated valuable time to advance our understanding about SARS-CoV-2 prevalence in healthcare workers. This study was also made possible by Prof. Jian Xu (Qingdao Institute of Bioenergy and Bioprocess Technology, Chinese Academy of Science), who generously donated nasopharyngeal swabs.

